# Critical Questions when Interpreting Coronavirus PCR Diagnostics

**DOI:** 10.1101/2020.06.11.20127241

**Authors:** Jürgen Durner, Siegfried Burggraf, Ludwig Czibere, Tobias Fleige, Michael Spannagel, David C Watts, Marc Becker

## Abstract

The results of PCR measurements are regarded as unquestionable. This statement must be put into perspective. This relativization is particularly important in connection with the interpretation of SARS-CoV-2 results. Members of the critical infrastructure, such as nurses, may be quarantined although this is not necessary and are therefore missing from patient care. With our small but impressive comparison of methods and transport media for SARS-CoV-2, we not only show the different sensitivity of common routine systems and media in laboratory medicine. Further, we would like to inform clinically working physicians, who are not familiar with the technical weaknesses of the PCR investigation, about gaps and present solutions for their daily work.

## Methodological Research

The outbreak of the new coronavirus SARS-CoV-2 is a significant challenge for the performance of biomedical analytical laboratories [1]. In the current situation with rapidly increasing demand for SARS-CoV-2 assays the focus is on the availability of test materials (smear material, test reagents) and test capacities. At present, there are essentially two indications for the use of PCR diagnostics. The first is the detection of the pathogen (or another cause of the current clinical symptoms), the second is the identification of possible contagiousness after the symptomatic phase of the disease. Especially in the second case the results of the PCR examination should be critically evaluated. In contrast to other PCR examinations, or laboratory medical analyses, currently SARS-CoV-2 diagnostic information about the device or the detection limit / sensitivity is not usually provided by the laboratory. Therefore, we investigated the influence of the type of transport medium and of two widely used commercial purification reagents on the sensitivity of SARS-CoV-2 detection. We compared these results with an automated detection system.

Since we did not have access to reference material with a defined number of copies, we have diluted one of our local positive patient samples. This hospitalized patient, showed typical symptoms and a positive PCR signal (i.e. a crossing point (Cp) value) similar to many other samples from patients in the acute phase of the disease (data not shown). To avoid possible fluctuations in sensitivity at low concentrations caused by the low amounts used in the tests, multiple determinations were carried out at a dilution of 10^−4^ in the experiments with different virus media. Using a protocol without purification of viral RNA, i.e. the *Munich Extraction Protocol* (**MEP**) [2], we could show that the type of transport medium had little influence on the detection sensitivity of SARS-CoV-2 in the PCR (Table 1).

**Table 1.**
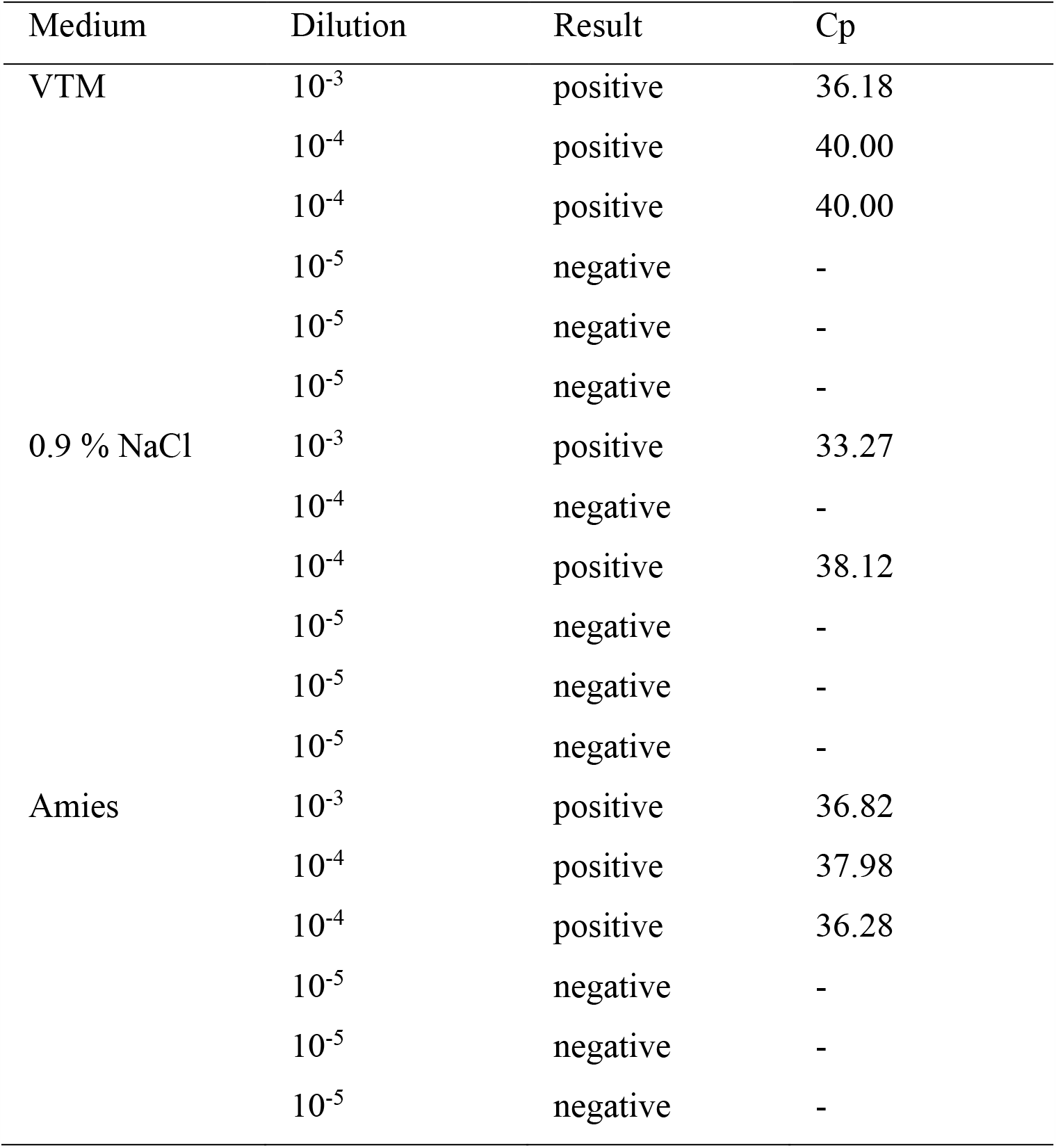
Comparison of different types of transport media for their influence on the detectability of SARS-CoV-2. VTM = virus transport medium; Cp = crossing point.

In Table 2 different purification methods are compared. Both purification systems showed the same sensitivity. Compared to the sensitivity when using **MEP**, the sensitivity when using commercially available preparation systems was one dilution level higher. This was mainly caused by concentration of the sample during the purification process that generated a higher input compared to the original sample in the PCR. When using the **MEP** it was not advisable to use increased volume in the test series of the different types of transport media, as these were directly inserted into the PCR without purification. This condition could introduce interfering substances.

**Table 2.**
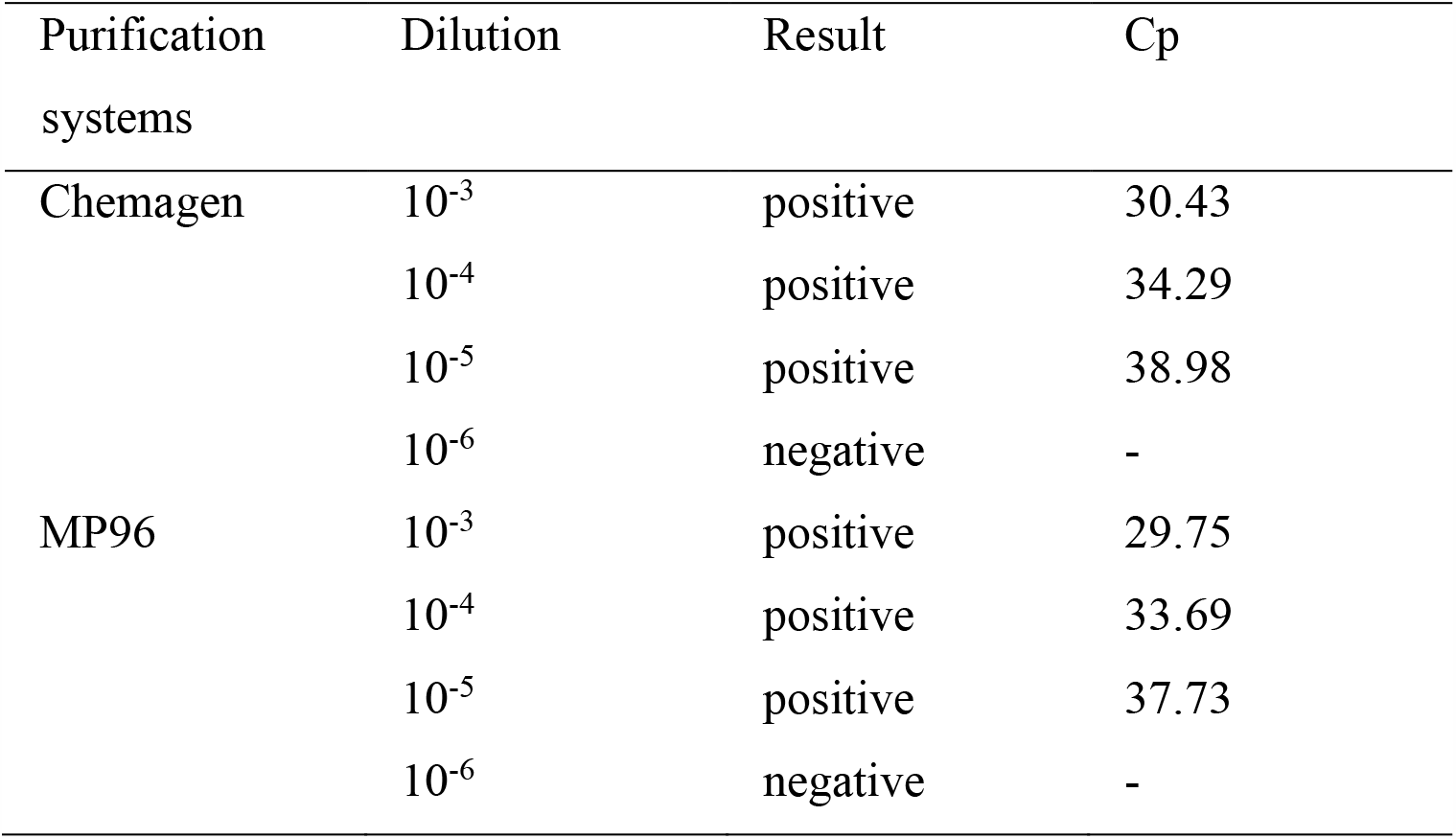
Comparison of different purification systems for their influence on the detectability of SARS-CoV-2 (Cp = crossing point).

Furthermore a fully automated system was tested. In contrast to normal PCR, this system uses two different primers set for two different targets (target 1 and target 2). By using this system, the sensitivity could be increased by at least one more dilution step compared to the use of commercial purification methods in PCR (Table 3). Again the main reason for the higher sensitivity was the higher sample input.

**Table 3.** Detectability of SARS-CoV-2 by using an automated diagnostic system (Ct = cycle threshold).

| automated diagnostic system | Dilution | Target 1 Result | Target 2 Result | Target 1 Ct | Target 2 Ct |
| --- | --- | --- | --- | --- | --- |
| Cobas 6800 | 10<sup>-5</sup> | positive | positive | 33.85 | 34.76 |
|  | 10<sup>-6</sup> | positive | positive | 34.11 | 35.83 |
|  | 10<sup>-7</sup> | positive | negative | 36.56 | - |
|  | 10<sup>-8</sup> | negative | negative | - | - |
|  | 10<sup>-9</sup> | negative | negative | - | - |

At level 10^−7^ we see the same phenomenon as in Table 1 at level 10^−4^. Due to the low viral load each primer was not amplified equally. Another possibility is that neither target had the same sensitivity.

An important result was apparent:

Close to the limit of detection, i.e. at dilution 10^−4^ (or 10^−7^ with the fully automated system) in our experiments it was possible to get both a positive and a negative result from the same sample. This is due to the fact that there is little RNA present due to the limit of detection. If a little more RNA molecules are present in one sample (via a suction effect through the pipette or by thermal influences), this may be sufficient to generate a small but measurable signal. Statistical thermodynamics provides another explanation. At high dilutions, RNA and primer molecules must find each other to react together. If this happens at the beginning of the PCR, the RNA is doubled in each PCR cycle. The earlier that this occurs, the more likely it is that a measurable signal is obtained at the end of the PCR.

This is clinically very relevant: It affects the question of whether or not the person has to be quarantined. For people working in the field of critical infrastructure, this means whether or not they are allowed to remain in active service. If not, this can result in a significant reduction in staff numbers, especially in hospitals.

How can we deal with this problem? In such cases there should be a close discussion with the clinically active colleagues. If clinical symptoms are present, it is recommended that the PCR examination be repeated by a new collection (“fresh sample”).

In addition, based on our experience and discussion with clinical colleagues, an active infection, combined with a high risk of infection of contact persons, usually results in such a high viral load that it can still be detected with **MEP** or the tested purification system. Based on data from our patient population, even a thousandfold (10^−3^) dilution was detected by all tested methods.

To digress: Does it make sense to test people by PCR who do not (or no longer) have symptoms? In these cases, the viral load may be at or below the detection limit of the chosen method. As we have also shown, multiple tests can sometimes show positive and sometimes negative values.

At this time, it cannot be clarified for SARS-CoV-2 whether or not there is a risk that can be eliminated of infection for contact persons despite PCR detection of the virus. It is known from other virus diseases that there is no longer any risk of infection after passing through the disease although the virus can be detected up to now [3]. In the case of SARS-CoV-2, it was shown that from the 8th day after the onset of symptoms, although viral RNA was still detectable in the throat, no infectious virus particles could be isolated [4]. This indicates that a “healing” of COVID-19 cannot be proven by PCR examination.

Does the analytical laboratory have a way of detecting borderline findings? In this context the Cp or Ct value (cycle threshold; both terms mean the same) is often cited. The Cp value indicates the number of cycles after the signal has been measured when the fluorescence signal from the exponential increase of the amplification of the virus is significantly above the background fluorescence for the first time. Cp values greater than 38 are considered an indication of lower amounts in the sample. It should be noted, however, that this value is only a *relative* measure of the concentration of the target in the PCR reaction, as it is influenced by many factors (reaction efficiency, method of determining the Cp value) [5]. Therefore Cp values from different PCR assays cannot be compared. The Cp value also depends on the pre-analysis (type of swab: gel, dry, transport medium). Due to the many factors influencing the Cp value, no recommendation should be made at this point as to whether or not it should be used for evaluation. If colleagues are unsure, it can only be recommended to consult the treating physician at this point and ask for the clinical situation of the patient.

It should be considered whether information on the detection limit / sensitivity should not be given so that a better classification of the significance of the result of the PCR measurement is possible.

The different sensitivities we have identified show that, although the lowest sensitivity was achieved with a simple purification reagent independent of the manufacturer a dilution up to ten thousand fold of the original sample was quite possible without falling below the detection limit. The question arises whether this is sufficient. It is not possible to define the necessary and clinically relevant sensitivity at this time. For the systems mentioned in Tables 2 and 3 there were delivery bottlenecks, so that delays occurred in analysis and thus in compilation of findings and patient evaluation / isolation.

The next step would be to consider quantification. Unfortunately, the necessary reference materials are not currently available for this. But even in this case, the clinical relevance of the number of copies for contagiousness should not be disregarded.

## Data Availability

All data was measured by us.

## Materials and Methods

### Comparison of different types of transport media

A stock solution was prepared for testing the sensitivity when using different transport media: virus transport medium (VTM; Yocon Biology Technology Company, Beijing, China, which was used in a commercially available swab) 0.9 % NaCl (Merck, Darmstadt, Germany) and Amies solution (MWE Medical Wire & Equipment, Crosham, United Kingdom). From a known positive SARS-CoV-2 sample 10 µl were taken and pipetted to 90 µl medium and different dilutions were prepared. The dilution steps were performed with the respective medium. From a dilution of 10^−4^ and lower, several PCR amplifications were performed from the dilutions.

Then, in a standard 96-well PCR plate (Nerbe plus, Winsen, Germany; preparation plate), 25 µl of water per well are distributed. Then 25 µl medium is pipetted into each well. The 96-well plate is then sealed with PCR foil (Nerbe plus) and heated to 92 °C for 10 min in a standard thermal cycler (Bio-Rad T100, Feldkirchen, Germany). The 96-well plate is then cooled to 12 °C and centrifuged for 2 min at 1000 rpm in a micro centrifuge (PlateFuge, Benchmark Scientific, Edison, New Jersey, USA).

One position each is used for the positive control and the negative control. The preparation was done according our Munich Extraction Protocol (MEP) [2].

### Comparison of purification systems

A stock solution was prepared for testing the sensitivity when using different purification systems: Chemagic Viral DNA/RNA 300 Kit H96 (Chemagen, PerkinElmer Chemagen Technologie, Baesweiler, Germany) and MagNA Pure 96 (MP96; Roche Diagnostics Mannheim, Germany). From a known positive SARS-CoV-2 sample 10 µl were taken and 90 µl of 0.9 % NaCl were pipetted and different dilution steps were prepared. The dilution steps were performed with 0.9 % NaCl. From a dilution of 10^−4^ and lower, several PCR amplifications were performed from the dilutions.

### Procedure of the PCR for the different types of transport media and the purification systems Preparation of PCR Master Mix

The master mix is produced from the RIDA®GENE SARS-CoV-2 RUO Kit (R-Biopharm, Darmstadt, Germany). 1380 µl reaction mix, 50 µl enzyme mix, and 70 µl internal control RNA are transferred into a 2 ml tube and mixed.

The preparation for this master mix is based on the manufacturer’s instructions. 15 µl of this PCR mix is distributed into each well of a qPCR 96-well plate (Roche Diagnostics).

### PCR Set up

Using the INTEGRA VIAFLO96 pipetting robot (INTEGRA Biosciences, Biebertal, Germany; 96 pipetting head), 5 µl of each sample is transferred simultaneously from the 96- well preparation plate to the 96-well qPCR containing the master. In addition, the positive control of the test kit is used, 5 µl water is used as negative control.

This qPCR plate is sealed with optical foil (Roche Diagnostics), centrifuged for 1 min (PCR PLATE SPINNER, VWR, Ismaning, Germany) and loaded into the LightCycler^®^ 480II (Roche Diagnostics).

### Amplification and Analysis

Realtime (RT)-PCR is performed using a 96 well block on a LightCycler^®^ 480II (Roche Diagnostics), with a reverse transcription step at 85 °C for 10 min, followed by denaturation at 95 °C for 1 min and by 45 cycles in two steps at 95 °C for 10 s and 60 °C for 15 s. A single acquisition of fluorescence signals is included in the 60 °C step. Detection of SARS-CoV-2 is at 510 nm and the internal amplification control at 580 nm.

### Automated diagnostic system

A stock solution was prepared to test the sensitivity when using an automated diagnostic system (Cobas^®^ 6800, Roche Diagnostics). From the known positive SARS-CoV-2 sample, 100 µl were taken and added to 900 µl 0.9 % NaCl to prepare the different dilution steps, which were then measured in the Cobas^®^ 6800.

## References

1. Binnicker MJ. Emergence of a Novel Coronavirus Disease (COVID-19) and the Importance of Diagnostic Testing: Why Partnership between Clinical Laboratories, Public Health Agencies, and Industry Is Essential to Control the Outbreak. Clin Chem. 2020. https://doi.org/10.1093/clinchem/hvaa071.

2. Durner J, Burggraf S, Czibere L, Fleige T, Madejska A, Watts DC, et al. Fast and simple high-throughput testing of COVID 19. Dent Mater. 2020. https://doi.org/10.1016/j.dental.2020.04.001.

3. Costantini VP, Cooper EM, Hardaker HL, Lee LE, Bierhoff M, Biggs C, et al. Epidemiologic, Virologic, and Host Genetic Factors of Norovirus Outbreaks in Longterm Care Facilities. Clin Infect Dis. 2016;62:1–10. https://doi.org/10.1093/cid/civ747.

4. Wolfel R, Corman VM, Guggemos W, Seilmaier M, Zange S, Muller MA, et al. Virological assessment of hospitalized patients with COVID-2019. Nature. 2020. https://doi.org/10.1038/s41586-020-2196-x.

5. Karlen Y, McNair A, Perseguers S, Mazza C, Mermod N. Statistical significance of quantitative PCR. BMC Bioinformatics. 2007;8:131. https://doi.org/10.1186/1471-2105-8-131.

